# Adaptation of ACMG/AMP guidelines for clinical classification of *BMPR2* variants in Pulmonary Arterial Hypertension resolves variants of unclear pathogenicity in ClinVar

**DOI:** 10.1101/2024.11.24.24317862

**Authors:** Christina A. Eichstaedt, Gabriel Maldonado-Velez, Rajiv D. Machado, Srimmitha Balachandar, Florence Coulet, Kristina Day, Dennis Dooijes, Melanie Eyries, Stefan Gräf, Daniela Macaya, Memoona Shaukat, Laura Southgate, Jair Tenorio-Castano, Wendy K. Chung, Carrie L. Welch, Micheala A. Aldred

**Affiliations:** Center for Pulmonary Hypertension, Thoraxklinik-Heidelberg gGmbH, at Heidelberg University Hospital and Translational Lung Research Center (TLRC), German Center for Lung Research (DZL), Heidelberg, Germany; Laboratory for Molecular Genetic Diagnostics, Institute of Human Genetics, Heidelberg University, Heidelberg, Germany; Division of Pulmonary, Critical Care, Sleep and Occupational Medicine, Department of Medicine, Indiana University School of Medicine, Indianapolis, IN, USA; Cardiovascular and Genomics Research Institute, City St George’s, University of London, School of Health & Medical Sciences, London, UK; Sorbonne Université, INSERM, Unité Mixte de Recherche Scientifique 938 et SIRIC CURAMUS, Centre de Recherche Saint-Antoine, Equipe Instabilité des Microsatellites et Cancer, Paris, France; UF d’Onco-angiogénétique et génomique des tumeurs solides, Département de Génétique, DMU BioGeM, GH Pitié-Salpêtrière, AP-HP, Sorbonne Université, Paris, France; Department of Genetics, University Medical Centre Utrecht, Utrecht University, Utrecht, the Netherlands; INSERM UMRS 1166, ICAN-Institute of CardioMetabolism and Nutrition, Sorbonne Université, Paris, France; Department of Medicine, School of Clinical Medicine, University of Cambridge, Victor Phillip Dahdaleh Heart & Lung Research Institute, Cambridge, UK; GeneDx, Inc, Gaithersburg, MD, USA; Institute of Medical and Molecular Genetics (INGEMM), Hospital Universitario La Paz, IDiPAZ, Universidad Autonoma de Madrid, Madrid, Spain; Centro de Investigación Biomédica en Red de Enfermedades Raras (CIBERER), Instituto de Salud Carlos III, Madrid, Spain; ITHACA, European Reference Network, Brussels, Belgium; Division of Genetics and Genomics, Department of Pediatrics, Boston Children’s Hospital and Harvard Medical School, Boston, MA, USA; Department of Pediatrics, Columbia University Irving Medical Center, New York, USA

**Keywords:** Pulmonary arterial hypertension, BMPR2, variant interpretation, ACMG/AMP guidelines, ClinVar

## Abstract

Purpose: Pulmonary arterial hypertension (PAH) is a rare disease that can be caused by pathogenic variants, most frequently in the bone morphogenetic protein receptor type 2 (*BMPR2*) gene. We formed a ClinGen variant curation expert panel to devise guidelines for the clinical interpretation of *BMPR2* variants identified in PAH patients. Methods: The general ACMG/AMP variant classification criteria were refined for PAH and adapted to *BMPR2* following ClinGen procedures. Subsequently, these specifications were tested independently by three members of the curation expert panel on 28 representative *BMPR2* variants selected from ClinVar, and then presented and discussed in the plenum. Results: Application of the final *BMPR2* variant specifications resolved 6 of 9 variants (66%) where multiple ClinVar classifications included a Variant of Uncertain Significance, with all six being reclassified as Benign or Likely Benign. Four splice site variants underwent clinically consequential reclassifications based on the presence or absence of supporting mRNA splicing data. Conclusion: The variant specifications provide an international framework and a useful tool for *BMPR2* variant classification and can be applied to increase confidence and consistency in *BMPR2* interpretation for diagnostic laboratories, clinical providers, and patients.

## INTRODUCTION

Pulmonary arterial hypertension (PAH) is a rare disease with a prevalence of 15-50 cases per one million individuals (Galiè et al. 2016; Humbert et al. 2023). Symptoms of PAH are non-specific including shortness of breath particularly during exertion, palpitations and, in advanced cases, edema, chest pain and dyspnea at rest. While non-invasive assessments such as echocardiography, electrocardiography, laboratory tests, pulmonary function tests and cardiopulmonary exercise testing can support a diagnosis of PAH, the definitive diagnosis is made by right heart catheterization. PAH is hemodynamically defined by an elevated mean pulmonary artery pressure >20 mmHg, pulmonary vascular resistance >2 Wood units and a pulmonary artery wedge pressure ≤15 mmHg (Humbert et al. 2023). PAH is currently classified as idiopathic (IPAH), heritable (HPAH), or associated with other diseases or exposures, including connective tissue disease, congenital heart disease, HIV infection or drugs and toxins. By definition, patients with HPAH have a positive family history and/or carry a pathogenic variant in a definitive PAH risk gene. Thus, apparently idiopathic patients may be reclassified as HPAH following genetic analysis.

The first gene identified as causative for PAH encodes the bone morphogenetic protein receptor type 2 (*BMPR2*) (Deng et al. 2000; Lane et al. 2000), and it remains the gene with the highest frequency of pathogenic variants in HPAH patients (Eichstaedt et al. 2023; Machado et al. 2015; Southgate et al. 2020). BMPR2 is a cell surface receptor and part of the transforming growth factor β superfamily responsible for vascular homeostasis of cell proliferation and apoptosis. Heterozygous Pathogenic or Likely Pathogenic *BMPR2* variants lead to PAH in an autosomal dominant manner with reduced penetrance of approximately 15-40% and show sexual dimorphism, with higher penetrance in females (Larkin et al. 2012).

Subsequently, several additional genes have been implicated in the aetiology of IPAH and HPAH, especially with the advent of next-generation sequencing of large cohorts (Gräf et al. 2018; Zhu et al. 2019; Zhu et al. 2021). A recent study carried out by our Clinical Genome Resource (ClinGen) pulmonary hypertension gene curation expert panel (PH GCEP), thoroughly characterized 27 genes and classified 12 of them to have a definitive gene-disease relationship with IPAH/HPAH (*ACVRL1*, *ATP13A3*, *BMPR2*, *CAV1*, *EIF2AK4*, *ENG*, *GDF2*, *KCNK3*, *KDR*, *SMAD9*, *SOX17*, *TBX4*), three to have moderate evidence (*ABCC8*, *GGCX*, *TET2*), six with limited evidence (*AQP1*, *BMP10*, *FBLN2*, *KLF2*, *KLK1*, *PDGFD*), and one to have no evidence for the relationship with PAH (*TOPBP1)* (Welch et al. 2023). Notably, four genes in the *BMPR2* pathway (*BMPR1A*, *BMPR1B*, *SMAD1*, *SMAD4*) were disputed as causative PAH genes (Welch et al. 2023).

ClinGen, funded by National Institutes of Health (NIH), serves as a central resource with information on the clinical significance of genes and variants (Rehm et al. 2015). Curation of genes and variants focuses on gene disease validity, variant pathogenicity, dosage sensitivity, and clinical actionability. ClinGen also offers the opportunity to form expert panels to curate not only genes but also variants (Rivera-Muñoz et al, 2018). To this end, following our gene curation efforts, we founded a PH variant curation expert panel (VCEP) to adapt the ACMG/AMP guidelines with a focus on *BMPR2*. We modified the general variant curation guidelines by the American College of Medical Genetics and Genomics (ACMG) and Association of Molecular Pathology (AMP) framework (Richards et al. 2015) for *BMPR2* variants specific to HPAH and/or IPAH. Since a significant number of *BMPR2* variants have an unclear impact on gene expression or function we aimed to resolve conflicting classifications to improve the interpretation of genetic test results. We tested our refined specification criteria on a pilot subset of 28 representative *BMPR2* variants, the results of which are described herein.

## METHODS

Within the framework of our Genetics Task Force of the International Consortium for Genetic Studies in PAH (PAH-ICON, https://pahicon.com/), we founded a ClinGen Pulmonary Hypertension VCEP with sixteen members from nine institutions across the USA and Europe, with representation from six countries (https://www.clinicalgenome.org/affiliation/50071/). Members included a chairperson (WKC), scientific lead/coordinator (CLW), expert reviewers, and biocurators. Expert reviewer and biocurator roles were assigned based on members’ experience but most members had dual roles. Seven members hold professional certifications in clinical genetics and/or regularly use ACMG/AMP guidelines to classify variants for clinical laboratory case sign-out. Most members had a history of ClinGen activities including curation of PAH gene relationships (Welch et al. 2023). All members received additional training in the ClinGen variant curation standard operating procedure v3, the 2015 ACMG/AMP guidelines, the ClinGen sequence variant interpretation working group recommendations for applying ACMG/AMP criteria, sequence variant literature searches, and use of ClinGen’s variant curation interface (VCI) (Preston et al. 2022). We followed the ClinGen four-step process (protocol version 9) to obtain VCEP approval (February 2024) with resulting variant classifications meeting U.S. Food and Drug Administration (FDA) recognition. A flow chart for the VCEP formation and activities is shown in Figure 1.

**Figure 1:**
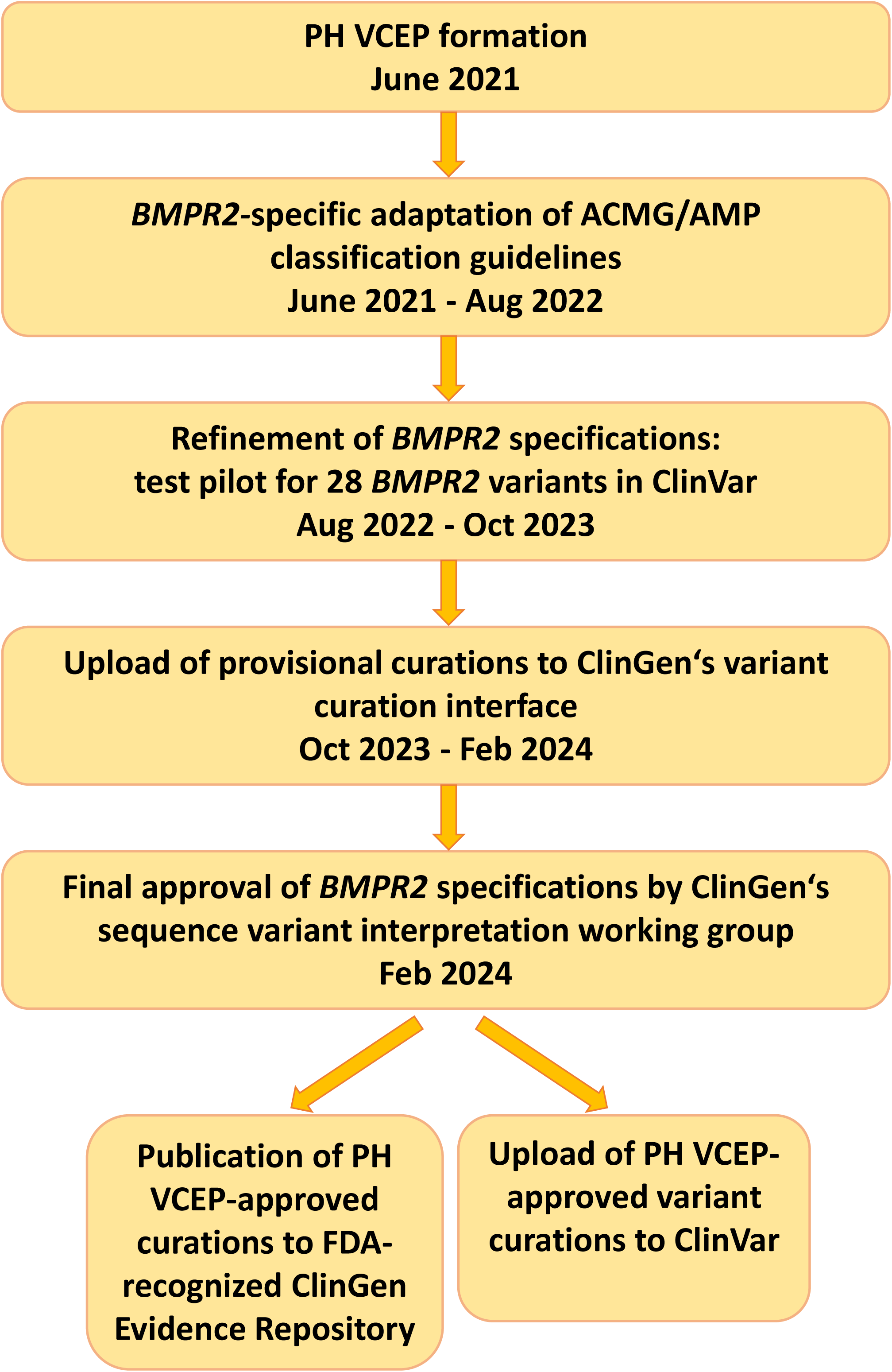
Flow chart of Pulmonary Hypertension Variant Curation Expert Panel formation and activities.

We limited the scope of this work to *BMPR2* variants because of the large number of loss-of-function variants, recurrent variants, and the relatively large body of experimental evidence defining critical domains/residues and functional effects of individual variants compared to other PAH risk genes (Welch et al. 2023). Monthly virtual meetings were held over a 14-month period (June 2021 to August 2022) to develop variant classification rules for *BMPR2* based on the ACMG/AMP guidelines (Figure 1). We then tested our adapted specifications on a pilot group of 28 variants, representing all ACMG classes, including those with conflicting interpretations of pathogenicity. The selected variants included insertions, deletions, 5’ untranslated region (UTR), missense, splice, nonsense, and synonymous variants. Only *BMPR2* variants identified in non-syndromic HPAH and/or IPAH cases were considered for pilot curation. Thus, one additional variant (c.319T>C p.Ser107Pro) was removed from the pilot curation list as it had been described in a proband with congenital heart disease associated PAH and was therefore outside the scope of this work. The *BMPR2* MANE Select transcript with accession number NM_001204.7 was used as the reference for all variant curations.

We assigned six variants to each curator and teams of three curators per variant. Provisional classifications were presented and discussed at monthly meetings over another 14-month period (August 2022 to October 2023). Modifications of our *BMPR2* criteria specifications were made over this period and following guidance of the ClinGen sequence variant interpretation working group. The specifications are available in the ClinGen Criteria Specification Registry (https://cspec.genome.network/cspec/ui/svi/doc/GN125). Final curations were uploaded to the VCI. Following final PH VCEP approval, variant curations were published to the ClinGen Evidence Repository and submitted to ClinVar (https://www.ncbi.nlm.nih.gov/clinvar/?term=bmpr2%5Bgene%5D&redir=gene). All members disclosed conflicts of interest and the curation assignments were mindful of conflicts. Our ACMG/AMP specifications are updated periodically. To find the most current information please visit https://cspec.genome.network.

## RESULTS

### 1. Development of *BMPR2*-specific variant curation criteria

#### Population frequency-based criteria: PM2, BA1, BS1 and BS2

The population frequency criteria evaluation includes four criteria (Table 1). Based on our *BMPR2* variant specifications, we applied PM2 as supporting when the variant was absent from the gnomAD v2.1.1 (controls) and v3.1.2 (controls/biobanks) populations or had an allele frequency below 0.01%. For this, we considered the ancestral subgroup with the highest frequency and at least 1,000 allele counts. The 0.01% threshold for allele frequency was defined based on the current prevalence of PAH of 15-50 cases/million individuals and accounting for reduced penetrance (Galiè et al. 2016; Humbert et al. 2023).

**Table 1:** Adapted *BMPR2* variant specifications for pulmonary arterial hypertension

| PATHOGENIC CRITERIA |  |  |
| --- | --- | --- |
| Criteria | Criteria Description | Specification |
| **VERY STRONG CRITERIA** |  |  |
| PVS1 | Null variant in a gene where loss of function is a known mechanism of disease. | Use *BMPR2*-specific PVS1 decision tree (Supplemental Fig.1) |
| **STRONG CRITERIA** |  |  |
| PS1 | Same amino acid change as a previously established pathogenic variant regardless of nucleotide change. | Downgrade to PS1_mod if variant is likely pathogenic. |
| PS2 | *De novo* (paternity confirmed) in a patient with the disease and no family history. | - |
| PS3 | Well-established *in vitro* or *in vivo* functional studies supportive of a damaging effect | If no validation controls (known pathogenic or benign variants), downgrade to PS3_supp. If the same functional assay has been performed for the same variant by an independent group and demonstrated to have the same functional effect by both groups, then PS3_strong. Can be applied additively with PP3. |
| PS4 | The prevalence of the variant in affected individuals is significantly increased compared with the prevalence in controls. | >4 probands: PS4_strong<br>>3 probands: PS4_mod<br>>1 proband PS4_supp<br>PM2 supporting must be met. Affected individual = mPAP>20 mmHg by right heart catheterization or estimated by echocardiography if RHC not advised. PS4 (at any strength) and PM2_supp can be used additively. |
| **MODERATE CRITERIA** |  |  |
| PM1 | Located in a mutational hot spot and/or critical and well-established functional domain. | Extracellular domain: p.Cys34, Cys60, Cys66, Cys84, Cys94, Cys99, Cys116, Cys117, Cys118, Cys123. Kinase domain: p.Gly210, Gly212, Lys230, Glu/Asn245, Asp333, Asn338, Asp351, Gly353, Glu386, Asp405, Gly410, Arg491. |
|  |  | Heterodimerization:<br>p.Asp485, Gln486,<br>Asp487, Ala488, Arg489,<br>Ala490, Arg491, Leu492. |
| PM2 | Absent/rare from controls in an ethnically-matched cohort population sample.<br>Note: We are deviating from “ethnically-matched cohort” because demographic/genetic ancestry data are not always available from ClinVar and other sources. | <0.01% among gnomAD controls, using the subpopulation with the highest frequency and at least 1,000 allele counts.<br>Always apply as PM2_supp.<br>PM2_supp and PS4 (at any strength) can be used additively. |
| PM3 | *For recessive disorders, detected in trans with a pathogenic variant.* | N/A |
| PM4 | Protein length changes due to in-frame deletions/insertions in a non-repeat region or stop-loss variants. | - |
| PM5 | Missense change at an amino acid residue where a different missense change determined to be pathogenic has been seen before. | If 2<sup>nd</sup> variant is pathogenic, then apply PM5_mod. If 2<sup>nd</sup> variant is likely pathogenic, then apply PM5_supp. |
| PM6 | Confirmed de novo without confirmation of paternity and maternity. | - |
| **SUPPORTING CRITERIA** |  |  |
| PP1 | Co-segregation with disease in multiple affected family members.<br>Note: at least 3 meioses/family required. | ≥7 meioses: PP1_strong<br>≥5 meioses: PP1_mod<br>≥3 meioses in a single family: PP1_supp |
| PP2 | *Missense variant in a gene that has a low rate of benign missense variation and where missense variants are a common mechanism of disease.* | N/A |
| PP3 | Multiple lines of computational evidence support a deleterious effect on the gene or gene product | REVEL ≥0.75 for missense, no up/downgrading;<br>SpliceAI ≥0.2 for non-canonical splice variants.<br>If no REVEL score or SpliceAI prediction available, then CADD ≥20 can be used. PP3 can be applied additively with PS3. |
| PP4 | *Phenotype specific for disease with single genetic etiology.* | N/A |
| PP5 | *Reputable source recently reports variant as pathogenic but the evidence is not available to the laboratory to perform an independent evaluation* | N/A<br>not to be used |
| **BENIGN CRITERIA** |  |  |
| **Criteria** | **Criteria Description** | **Specification** |
| **STAND ALONE CRITERIA** |  |  |
| BA1 | Allele frequency above X% | 1% |
| **STRONG CRITERIA** |  |  |
| BS1 | Allele frequency greater than expected for disease (>X%)<br>Note: PAH prevalence: 15-50 cases/million individuals | ≥0.1% among gnomAD controls, using the subpopulation with the highest frequency and at least 1,000 allele counts. |
| BS2 | Observed in the homozygous state in a healthy adult | ≥3 counts: BS2_strong; ≥2 counts: BS2_supp |
| BS3 | Well-established *in vitro* or *in vivo* functional studies shows no damaging effect on protein function | BMP2 functional assay documentation with appropriate controls |
| BS4 | Lack of segregation in affected members of a family. | Lack of disease segregation among variant carriers should not be scored due to low penetrance of PAH variants in families. Lack of variant segregation in affected family members should be scored. |
| **SUPPORTING CRITERIA** |  |  |
| BP1 | *Missense variant in gene where only loss-of-function causes disease* | N/A |
| BP2 | *Observed in trans with a pathogenic variant for a fully penetrant dominant gene/disorder; or observed in cis with a pathogenic variant in any inheritance pattern.* | N/A |
| BP3 | In-frame deletions/insertions in a repetitive region without a known function | - |
| BP4 | Multiple lines of computational evidence suggest no impact on gene or gene product | REVEL ≤0.25, no up/downgrading. SpliceAI ≤0.1 for non-canonical splice variants. If no REVEL score or SpliceAI prediction available, then CADD ≤10 can be used. |
| BP5 | *Variant found in a case with an alternate molecular basis for disease* | N/A |
| BP6 | *Reputable source recently reports variant as benign but the evidence is not available to the laboratory to perform an independent evaluation* | N/A<br>not to be used |
| BP7 | A synonymous (silent) variant for which splicing prediction algorithms predict no impact to the splice consensus sequence nor the creation of a new splice site AND the nucleotide is not highly conserved. | - |

mod, moderate; supp, supporting; PAH, pulmonary arterial hypertension; VUS, variant of uncertain significance.

Similar to PM2, we considered the highest allele frequency among ancestral subgroups on gnomAD control datasets to apply BA1 as a stand-alone criterion for benign variants with an allele frequency >1 % or ≥0.1% for BS1, an allele frequency which was greater than expected for PAH in controls. The presence of homozygotes was scored with BS2 as supporting for ≥2 controls and as strong for ≥3 controls.

#### Relevant variant frequency in patients: PS4, PP1 and BS4

Various criteria address the variant frequency in related or unrelated H/IPAH patients. While disease causing *BMPR2* variants are often novel, a recurrence in unrelated individuals can be used as evidence for criterion PS4. If a variant has a very low population frequency (PM2, see above) and the same variant has been identified previously in >4 unrelated H/IPAH patients, PS4_strong can be applied (Table 1). The criterion is moderate for >3 previously described cases and supporting for >1. Recurrent variants were identified through ClinVar, together with unpublished data from the labs of VCEP members and a thorough literature search for published variants not deposited in ClinVar. Importantly, care was taken to avoid counting duplicate reports of the same or related individuals across different publications. Also, co-segregation with disease lends strength to a variant. Analogous to the cardiomyopathy working group (Kelly et al. 2018), we used PP1 as strong with ≥7 meioses, moderate for ≥5 meioses across or within families and supporting for ≥3 meioses within a single family. In contrast, the lack of segregation of a variant with the disease in an affected PAH patient can be used as benign evidence for BS4. Due to the reduced penetrance of *BMPR2* variants, their presence in unaffected family members was not considered as evidence for benignity.

#### Null variant decision tree: PVS1

The criterion with the strongest evidence for pathogenicity is PVS1 (Table 1). The general PVS1 decision tree (Abou Tayoun et al. 2018) was adapted for *BMPR2* (see Supplementary Figure 1). The criterion applies to predicted null variants leading to a loss of function. Thus, it mainly refers to nonsense variants introducing a premature stop codon or frameshift variants and canonical splice site variants (+1,+2,-1,-2) likely inducing nonsense mediated decay (NMD). For such variants affecting *BMPR2* exons 1 to 11 and all but the final 50 nucleotides of penultimate exon 12 (c.1_c.2816), “PVS1_very strong” could be applied. A scoring as “strong” was possible if splice site variants were located at the first or second intronic base pair and were predicted to result in exon skipping, leading to an in-frame transcript with a loss or truncation of the ligand domain p.33_131, the transmembrane domain p.151_171, the kinase domain p.203_504 or the heterodimerization domain p.485_492.

#### Functional assay characterisation: PS3 and BS3

The PS3 criterion involves *in vitro* or *in vivo* functional studies supportive of a damaging effect on the *BMPR2* gene or protein according to the ACMG guidelines. To determine whether a variant met the PS3 criterion, we considered quantitative and qualitative assays, as described below. In order to be considered as valid evidence, quantitative assays needed to include biological and technical replicates, positive controls (wild-type *BMPR2*/BMPR2, endogenous or total protein), validation controls (previously established, known Pathogenic or Benign variants within the same assay), negative controls (empty vectors), and robust statistical analysis (usually *t* test or ANOVA). Qualitative cytoplasmic retention assays did not require statistical analysis to apply PS3 as long as they included appropriate controls and replicates. Functional studies that failed to include validation controls could apply PS3 at the supporting level (Brnich et al. 2019) at the discretion of the curator and PH VCEP.

#### Gene reporter/luciferase assays

Luciferase assays may be employed to investigate the functional impact of missense variants. Typically, cells are transfected with a combination of plasmids containing i) BMP-responsive element from a SMAD promoter incorporated with a luciferase (Lux) gene; ii) wild-type and mutant BMPR2, e.g. introduced by site-directed mutagenesis; iii) cognate type I receptors (ALK1/2/3/6). Luciferase activity is then measured, normalised to beta-galactosidase or alkaline phosphatase activity. When compared to wild-type BMPR2, these analyses either confirmed abrogation of SMAD-mediated signalling activity for previously established kinase-dead variants or identified variants that retained enzymatic activity with a luciferase signal comparable to wild-type (Nasim et al. 2008; Nishihara et al. 2002).

#### Cell proliferation assays

The gold standard for assessing changes in cell proliferation is to quantify the percentage of DNA-synthesizing cells. This can be accomplished by measuring incorporation of thymidine, or thymidine analogs bromodeoxyuridine (BrdU) or 5-ethynyl-2’-deoxyuridine (EdU), into *de novo* DNA. In PAH, proliferation of human pulmonary arterial smooth muscle, endothelial, or microvascular endothelial cells from H/IPAH transplant patients is compared to unaffected controls (i.e. unused donor tissue). Proliferation of similar cell types from mice heterozygous for patient-specific *Bmpr2* variants can be compared to wild-type (Long et al. 2006). Growth curve proliferation assays involving direct cell counting are often included as an independent assessment of cell proliferation (Long et al. 2006; Morrell et al. 2001). Basic controls should include full growth medium with 10% fetal bovine serum (FBS) (positive control) and serum restricted medium with 0.1% FBS (negative).

#### Protein binding assays

Protein binding assays are designed to investigate interaction of a protein with another protein, ligand-receptor binding, or transient protein binding for signal transduction. The acceptable protein assays in our PH VCEP panel were qualitative immunoprecipitation, glutathione-S-transferase (GST) pull-down and quantitative radioligand binding assays (Foletta et al. 2003; Machado et al. 2003; Nasim et al. 2008; Nishihara et al. 2002; Rudarakanchana et al. 2002; Zakrzewicz et al. 2007; Zhang et al. 2003). We considered confirmed ligands (bone morphogenetic proteins, BMPs) or coreceptors (activin receptors, ALKs) as acceptable binding partners while unconfirmed ligands or receptors were not scored. Variant location within BMPR2 domains e.g. ligand binding domain or kinase domain were also taken into consideration during assessment of protein binding assays (Nasim et al. 2008; Rudarakanchana et al. 2002; Zakrzewicz et al. 2007). In addition, downstream effector readouts were limited to SMADs and p38 while any unconfirmed molecules were not assessed. Positive control of wild-type BMPR2 and negative control of empty vector were considered for upgrading or downgrading the functional criteria.

#### SMAD phosphorylation assays

For SMAD phosphorylation assays, the ability of a BMPR2 variant to phosphorylate SMAD proteins should be compared to controls, preferably unphosphorylated total SMAD proteins or alternatively, housekeeping genes on a Western blot with densitometry. Negative controls were either cells without transfected BMPR2, with BMPR2 wild-type, or cells from non-PAH controls. Variants that demonstrated no deleterious effect with levels comparable to wild-type BMPR2 were scored under BS3, or BS3_supporting if validation controls were lacking.

#### Definition of critical domains: PM1

For the PM1 criterion, critical BMPR2 functional domains were defined as the extracellular or ligand binding domain and serine-threonine kinase domain, containing a heterodimerization motif, separated by a transmembrane domain. These domains are delineated by strong evidence from structural studies of the activin type II receptor extracellular domain, evolutionary conservation across eukaryotic kinases and hydrogen deuterium exchange mass spectrometry analysis of the interface between BMPR2 and the ALK2 kinase domain. Moreover, gene reporter assays have indicated the relative importance of amino acid residues within these regions (Agnew et al. 2021; Greenwald et al. 1999; Hanks and Hunter 1995; Nasim et al. 2008; Nishihara et al. 2002). Ten cysteine residues within the extracellular domain are required for formation of five disulphide bonds essential for correct three-dimensional folding (p.Cys34, p.Cys60, p.Cys66, p.Cys84, p.Cys94, p.Cys99, p.Cys116, p.Cys117, p.Cys118 and p.Cys123). Sequences from 60 aligned eukaryotic kinases indicate the presence of eight evolutionarily invariant residues (p.Gly212, p.Lys230, p.Asp333, p.Asn338, p.Asp351, p.Glu386, p.Asp405 and p.Arg491) and four nearly invariant residues (p.Gly210, p.Glu/Asn245, p.Gly353, p.Gly410). Moreover, structural analyses demonstrated that BMPR2/ALK heterodimerization is contingent on amino acids p.Asp485_Leu492 (Agnew et al. 2021; Greenwald et al. 1999; Hanks and Hunter 1995). Variants located at these critical amino acid residues were upgraded to PM1_strong. The transmembrane domain was considered a critical domain for loss of function variants (nonsense/frameshift/splice site) involving exon 4 but not for missense variants.

#### Minor adaptations: PM5 and PS1

In this study, PM5 at the moderate level was applied when a missense variant was found in the same amino acid as a previously described Pathogenic missense variant. If the previous missense variant was classified as Likely Pathogenic, then PM5 was applied at the supporting level. PS1 could be applied as strong, if the same amino acid substitution was described previously as Pathogenic and as moderate if the variant was considered Likely Pathogenic. Alternative variants were curated using our PH VCEP specifications rather than relying on existing interpretations published in ClinVar or elsewhere.

#### *In silico* tools: PP3/BP4

The PP3 and BP4 criteria consider the computational evidence that supports the presence (PP3) or absence (BP4) of a predicted, deleterious effect on *BMPR2*. Our VCEP adopted the rare exome variant ensemble learner (REVEL) (Ioannidis et al. 2016), Splice AI (Jaganathan et al. 2019), and Combined Annotation Dependent Depletion (CADD) (Kircher et al. 2014) scores to apply PP3 (REVEL ≥0.75; Splice AI ≥0.2) or BP4 (REVEL ≤0.25; Splice AI ≤0.1). In cases when REVEL or Splice AI scores were not available, a CADD ≥20 (PP3) or ≤10 (BP4) was considered.

Specifications for all criteria are detailed in Table 1 and Supplementary Figure 1. The *de novo* criteria (PS2 and PM6), deletion/insertion change criteria (PM4 and BP3) and the criterion for synonymous variants without splice site effects (BP7) were applied unaltered. PM3, PP2, PP4, BP1, BP2 and BP5 were considered not applicable to *BMPR2*-related PAH. Criteria referring to a “reputable source” (PP5 and BP6) are no longer used according to ClinGen revised guidance (Biesecker et al. 2018). Criteria were combined using the Bayesian points scoring system outlined by Tavtigian et al. 2020 (Tavtigian et al. 2020) with a modification for Likely Benign, as follows: Pathogenic (P), ≥10 points; Likely Pathogenic (LP), 6 to 9; Variant of Uncertain Significance (VUS), -1 to 5; Likely Benign (LB), -2 to -6; Benign (B), ≤ -7.

### 2. Pilot testing of PH VCEP specifications

The results of our pilot curation of 28 *BMPR2* variants are detailed in Table 2 and Figure 2. The classification of 7 variants (25%; 6 Pathogenic and 1 VUS) did not change from the existing ClinVar classification. These included well-established Pathogenic variants that were included as “positive controls” to test our specifications. If the P/LP and B/LB classifications are grouped per their clinical utility, a further 7 variants were unchanged, including 4 with mixed B/LB classifications in ClinVar. Three of these were resolved to Likely Benign and one to Benign when our specifications were applied, although this is not consequential for their clinical interpretation. Of the remaining 14 variants (50%), 9 had discrepant classifications in ClinVar. Eight of these were VUS/LB or VUS/LB/B, of which 3 were resolved to Benign, 3 to Likely Benign, and 2 remained as VUS. Two variants changed from LB to VUS or vice versa. The most pronounced and clinically consequential changes were seen with intronic splice region variants, with 3 variants (c.247+1_247+7del, c.529+2T>C and c.968-3C>G) reclassified from Pathogenic or P/VUS to VUS, and 1 from VUS to Pathogenic (c.968-5A>G). As discussed further below, mRNA analysis to confirm the predicted consequences of splice site variants is critical for accurate classification, especially for variants outside of the canonical positions.

**Table 2:** Detail of the *BMPR2* variants curated in the pilot study

| **Variant\*** | **Variant type** | **ClinVar classification** | **PH VCEP classification** | **ACMG/AMP criteria and points applied** | **Score<sup>†</sup>** |
| --- | --- | --- | --- | --- | --- |
| c.-924A>G | 5' untranslated region | VUS/Likely Benign | Benign | BA1 -8, BS2_supp -1, BP4 -1 | -10 |
| c.-669G>A | 5' untranslated region | Likely Benign/ Benign | Benign | BA1 -8 | -8 |
| c.218C>G (p.Ser73*) | nonsense | Pathogenic | Pathogenic | PVS1 +8, PS4_supp +1, PM2_supp +1 | +10 |
| c.240_241insT (p.Lys81*) | nonsense | Pathogenic | Likely Pathogenic | PVS1 +8, PM2_supp +1 | +9 |
| c.247+1_247+7del | splice region / intronic | Pathogenic | VUS | PVS1_strong +4, PM2_supp +1 | +5 |
| c.251G>A (p.Cys84Tyr) | missense | Pathogenic | Pathogenic | PS4_mod +2, PM1_strong +4, PM2_supp +1, PM5 +2, PP3 +1 | +10 |
| c.354T>G (p.Cys118Trp) | missense | Pathogenic (3) | Pathogenic | PS3 +4, PS4 +4, PM1_strong +4, PM2_supp +1, PM5_supp +1, PP1 +1, PP3 +1 | +16 |
| c.419-38del | splice region / intronic | VUS/Likely Benign/Benign | Benign | BA1 -8, BS2 -4, BP4 -1 | -13 |
| c.529+2T>C | canonical splice site | Pathogenic | VUS | PVS1_strong +4, PM2_supp +1 | +5 |
| c.545G>A (p.Gly182Asp) | missense | VUS (3)/Likely Benign (1) | Likely Benign | BS3 -4, PP3 +1 | -3 |
| c.797G>C (p.Arg266Thr) | missense | VUS | VUS | PM2_supp +1 | +1 |
| c.901T>C (p.Ser301Pro) | missense | Likely Pathogenic | Pathogenic | PS3 +4, PS4 +4, PM1 +2, PM2_supp +1, PM6 +2 | +13 |
| c.968-5A>G | splice site / intronic | VUS (3) | Pathogenic | PVS1 +8, PS4_supp +1, PM2_supp +1, PP3_supp +1 | +11 |
| c.968-3C>G | splice site / intronic | Pathogenic/VUS | VUS | PS4_supp +1, PM2_supp +1, PP3 +1 | +3 |
| c.1040G>A (p.Cys347Tyr) | missense | Pathogenic | Likely Pathogenic | PS3_supp +1, PS4 +4, PM2_supp +1, | +8 |
|  |  |  |  | PM5_supp +1,<br>PP3 +1 |  |
| c.1042G>A<br>(p.Val348Ile) | missense | Likely Benign/<br>Benign | Likely<br>Benign | BS1 -4, PS3_supp<br>+1 | -3 |
| c.1413+1G>A | canonical splice<br>site | Pathogenic | Pathogenic | PVS1 +8, PS4_mod<br>+2, PM2_supp +1 | +11 |
| c.1424C>A<br>(p.Ser475*) | nonsense | Pathogenic (2) | Pathogenic | PVS1 +8,<br>PS4_supp +1,<br>PM2_supp +1 | +10 |
| c.1472G>A<br>(p.Arg491Gln) | missense | Pathogenic | Pathogenic | PS2 +4, PS3_supp<br>+1, PS4 +4,<br>PM1_strong +4,<br>PM2_supp +1,<br>PM5 +2, PP1 +1,<br>PP3 +1 | +18 |
| c.1481C>T<br>(p.Ala494Val) | missense | Likely Benign | VUS | PM1 +2, PP3 +1,<br>BS1 -4 | -1 |
| c.1509A>C<br>(p.Glu503Asp) | missense | VUS | Likely<br>Benign | BS3 -4 | -4 |
| c.1698T>A<br>(p.Ile566=) | synonymous | VUS/Likely<br>Benign | VUS | PM2_supp +1, BP4<br>-1, BP7 -1 | -1 |
| c.1766A>G<br>(p.Tyr589Cys) | missense | Likely Benign/<br>Benign | Likely<br>Benign | BS1 -4 | -4 |
| c.2186G>C<br>(p.Gly729Ala) | missense | VUS/Likely<br>Benign | Likely<br>Benign | BS1 -4 | -4 |
| c.2352C>T<br>(p.Val784=) | synonymous | VUS/Likely<br>Benign | VUS | PM2_supp +1, BP7<br>-1 | 0 |
| c.2618G>A<br>(p.Arg873Gln) | missense | VUS/Likely<br>Benign | Benign | BS1 -4, BS3_supp -<br>1, BS4 -4 | -9 |
| c.2887G>T<br>(p.Gly963Cys) | missense | Likely Benign/<br>Benign | Likely<br>Benign | BS1 -4 | -4 |
| c.2948G>A<br>(p.Arg983Gln) | missense | VUS (3)/Likely<br>Benign (1)<br>/Benign (1) | Likely<br>Benign | BS1 -4 | -4 |

\*Variant nomenclature refers to transcript NM\_001204.7; †scoring modified from Tavtigian et al. 2020: pathogenic (P), ≥10 points; likely pathogenic (LP), 6 to 9; VUS, -1 to 5; likely benign (LB), -2 to -6; benign (B), ≤ -7; mod, moderate; supp, supporting; VUS, variant of uncertain significance.

**Figure 2:**
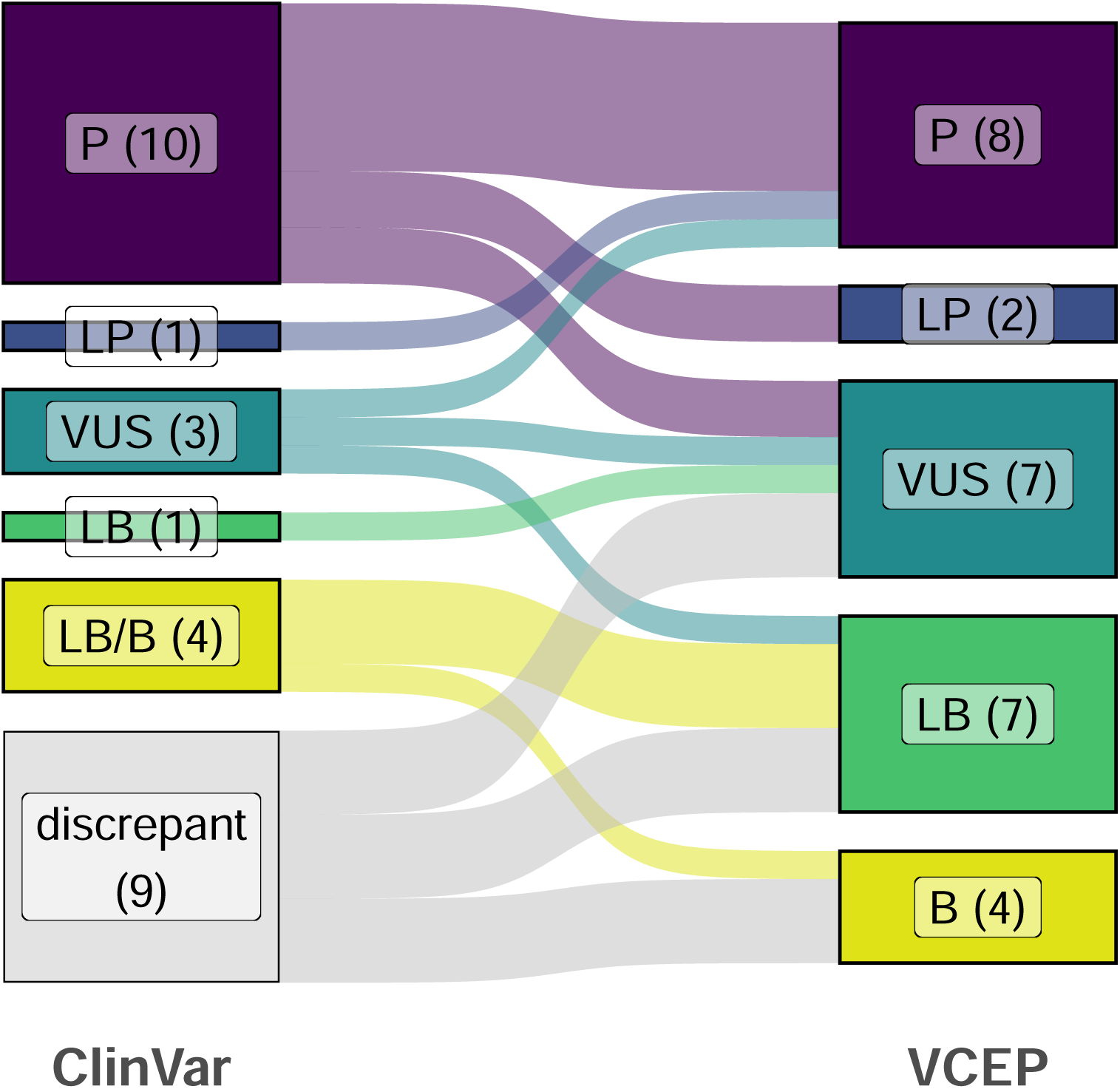
Comparison of ClinVar and PH VCEP classifications. Sankey plot shows the effect of applying our VCEP variant specifications (right) compared with the original ClinVar classification(s) (left) for the pilot panel of 28 *BMPR2* variants. P, pathogenic; LP, likely pathogenic; VUS, variant of uncertain significance; LB, likely benign; B, benign.

### 3. Evaluation of newer bioinformatics tools: AlphaMissense and BayesDel

Lastly, we performed a pilot evaluation of two newer bioinformatic prediction tools not covered in the current ACMG/AMP guidelines. As a novel and emerging *in silico* prediction tool for missense variants, AlphaMissense, developed by Google DeepMind, calculates missense variant pathogenicity by combining structural context and evolutionary conservation, using data from AlphaFold prediction and human and primate variant frequency databases (Cheng et al. 2023). It generates scores between 0 and 1, with higher scores predicting greater likelihood of pathogenicity. Suggested thresholds for likely pathogenic and likely benign are >0.564 and <0.340, respectively. BayesDel (no AF) is a deleteriousness meta-score. The range of the score is from -1.29334 to 0.75731. The higher the score, the more likely the variant is pathogenic (Pejaver et al. 2022). Both programmes were applied in addition to REVEL and CADD to the 15 missense variants in our pilot trial and compared to our final VCEP classification. AlphaMissense outperformed REVEL, CADD and BayesDel by reaching an agreement with our experts for 12/15 variants while the other three programmes each only supported 5/15 assessments (Table 3).

**Table 3:** Comparison of three *in silico* prediction programmes with our VCEP classification of missense variants

| Variant* | PH VCEP classification | REVEL | CADD | AlphaMissense | BayesDel |
| --- | --- | --- | --- | --- | --- |
| c.251G>A<br>(p.Cys84Tyr) | Pathogenic | Deleterious (0.95) | Deleterious (32) | Likely Pathogenic (0.998) | Deleterious (Strong) (0.59) |
| c.354T>G<br>(p.Cys118Trp) | Pathogenic | Deleterious (0.93) | Deleterious (28.5) | Likely Pathogenic (0.999) | Deleterious (Strong) (0.53) |
| c.545G>A<br>(p.Gly182Asp) | Likely Benign | Deleterious (0.81) | Deleterious (28.0) | Uncertain (0.425) | Deleterious (Moderate) (0.36) |
| c.797G>C<br>(p.Arg266Thr) | VUS | Uncertain (0.63) | Deleterious (29.5) | Likely Pathogenic (0.893) | Deleterious (Supporting) (0.21) |
| c.901T>C<br>(p.Ser301Pro) | Pathogenic | Uncertain (0.45) | Deleterious (25.1) | Likely Pathogenic (0.905) | Deleterious (Supporting) (0.16) |
| c.1040G>A<br>(p.Cys347Tyr) | Likely Pathogenic | Deleterious (0.94) | Deleterious (31) | Likely Pathogenic (0.994) | Deleterious (Moderate) (0.48) |
| c.1042G>A<br>(p.Val348Ile) | Likely Benign | Uncertain (0.74) | Deleterious (26.2) | Likely Benign (0.201) | Deleterious (Moderate) (0.39) |
| c.1472G>A<br>(p.Arg491Gln) | Pathogenic | Deleterious (Strong) (0.96) | Deleterious (34) | Likely Pathogenic (0.992) | Deleterious (Strong) (0.62) |
| c.1481C>T<br>(p.Ala494Val) | VUS | Deleterious (Moderate) (0.87) | Deleterious (31) | Likely Pathogenic (0.979) | Deleterious (Moderate) (0.4) |
| c.1509A>C<br>(p.Glu503Asp) | Likely Benign | Uncertain (0.66) | Uncertain (17.7) | Likely Benign (0.26) | Deleterious (Moderate) (0.29) |
| c.1766A>G<br>(p.Tyr589Cys) | Likely Benign | Uncertain (0.58) | Deleterious (28) | Likely Benign (0.263) | Deleterious (Supporting) (0.18) |
| c.2186G>C<br>(p.Gly729Ala) | Likely Benign | Uncertain (0.36) | Deleterious (23.6) | Likely Benign (0.101) | Uncertain (0.06) |
| c.2618G>A<br>(p.Arg873Gln) | Benign | Uncertain (0.55) | Deleterious (26) | Likely Benign (0.266) | Deleterious (Moderate) (0.34) |
| c.2887G>T<br>(p.Gly963Cys) | Likely Benign | Uncertain (0.42) | Deleterious (26.8) | Likely Benign (0.099) | Uncertain (-0.04) |
| c.2948G>A<br>(p.Arg983Gln) | Likely Benign | Uncertain (0.4) | Deleterious (24.8) | Likely Benign (0.321) | Uncertain (-0.04) |
| **Agreement with VCEP** | - | 5/15 | 5/15 | 12/15 | 5/15 |

\*Variant nomenclature refers to transcript NM\_001204.7
AlphaMissense: likely pathogenic, 0.564-1.0; uncertain, 0.34-0.564; likely benign, 0-0.34.
BayesDel: $\geq 0.5$ : strong pathogenic evidence, 0.27-0.5: moderate pathogenic, 0.13-0.27: supporting pathogenic, -0.36 to -0.18: supporting benign, and score $\leq -0.36$ moderate benign; (Pejaver et al. 2022)
CADD: $\geq 20$ pathogenic, 10-20 uncertain, $\leq 10$ benign
REVEL: $\geq 0.75$ pathogenic, 0.25-0.75 uncertain, $\leq 0.25$ benign
VUS: variant of uncertain significance

## DISCUSSION

We adapted the general ACMG/AMP variant specification criteria specifically to *BMPR2*, the gene most commonly implicated in PAH, with loss or reduced function as the mechanism of action. This guidance should be valuable to improve harmonization in reporting variants identified in patients with heritable or idiopathic PAH across clinical laboratories. Of note, we did not consider variants identified in patients with associated PAH, therefore these specifications may require modification in such cases.

Unlike null variants such as canonical splice site or frameshift variants, it is often challenging to classify novel and rare missense variants as Likely Pathogenic/Pathogenic with currently available data. In many cases, functional data are lacking and previous reports of the same variant in other PAH patients are usually limited. This often leaves only PM2 (low population frequency) and PP3 (pathogenicity predictions) to support missense variant interpretation. Hence, it is crucial for patients and clinicians that all variants are uploaded to ClinVar to potentially identify recurrent variants or amino acid changes to apply additional criteria (PS4, PS1, PM5). Accurate predictive testing for the familial disease-causing variant is only possible for Likely Pathogenic or Pathogenic variants, and genetic risk prediction is critical as there are increasingly effective therapies for PAH that are likely more effective when applied earlier in the disease course.

Of the 28 *BMPR2* variants curated in this pilot study, only seven (25%) had functional analyses that could be scored, of which five provided evidence for PS3 and two for BS3. Access to tissues, cell lines, and nucleic acids to perform the functional analyses considered in this study is not always possible and, when available, the materials and resources may be limited. The lack of functional analyses presents challenges when determining the impact of a given reported genomic variant. In this regard, our pilot testing of AI-based prediction algorithms, including AlphaMissense, suggests that such programs may be highly accurate in predicting the pathogenicity of missense variants in the absence of functional data. Furthermore, the assessment of mRNA in patients with suspected splice site variants located more than two base pairs within the intron is relatively straightforward and provides a key experimental indication of pathogenicity. The importance of this is highlighted by three splice site variants in our pilot panel, including one at the +2 position of a splice donor site, that were downgraded from Pathogenic to VUS due to the lack of supporting mRNA data. Conversely, one variant in the -5 splice acceptor position was upgraded from VUS to Pathogenic on the basis of mRNA analysis performed in the lab of a VCEP member that provided evidence for PVS1.

In this study, we encountered a large Iberian family in which 32 individuals were genotyped and 22 were heterozygous for c.1472G>A p.Arg491Gln co-segregating with PAH (Puigdevall et al. 2019). However, segregation analyses are typically challenging due to small family sizes, the work needed to organize family studies, and incomplete penetrance. Due to the need for cardiac catheterization for clinical diagnosis, pulmonary hemodynamics may not be available for some family members. In addition, without enough affected heterozygotes, the number of meioses observed may not be sufficient to use the segregation criteria PP1 or BS4. The presence or absence of this evidence can significantly influence the final classification of a given variant. Therefore, it is essential to develop protocols or policies to collect relevant data from large families when there is suspicion of HPAH. Moreover, clinical guidelines recommend offering cascade testing to potentially identify at-risk heterozygotes and offer clinical screening to diagnose PAH as early as possible (Eichstaedt et al. 2023).

In conclusion, these newly adapted ClinGen specification guidelines to assess *BMPR2* variants for PAH patients will allow more reliable and accurate variant classification. Although the overall proportion of VUS increased, this reflects that approximately one-third of the chosen variants had discrepant classifications in ClinVar. Notably, we were able to resolve 6 of the 8 variants with a discrepant VUS/LB, reclassifying them as Benign or Likely Benign. The remaining VUS may be reclassified with increasing evidence in the future. Benign or Likely Benign variants are most likely unrelated to the disease and thus do not increase the genetic risk of developing PAH. In contrast, Pathogenic or Likely Pathogenic variants allow for risk stratification of other family members. Thus, the consensus specification document substantially aids in providing patients and caregivers with an accurate genetic diagnosis which may play an increasing role in therapy.

### Author contributions

Conceptualization, methodology and data curation: all authors

Supervision: CLW, WKC

Writing – original draft: CAE, GMV, RDM, SG, MS, CLW, MAA

Writing – review and editing: all authors.

**Ethics statement:** all data were obtained from ClinVar and/or the published literature, and were fully de-identified. Therefore, IRB approval and informed consent was not required.

## Data Availability

The BMPR2 specifications are available in the ClinGen Criteria Specification Registry (https://cspec.genome.network/cspec/ui/svi/doc/GN125). Variant curations were published to the ClinGen Evidence Repository and submitted to ClinVar (https://www.ncbi.nlm.nih.gov/clinvar/?term=bmpr2%5Bgene%5D&redir=gene).

https://www.ncbi.nlm.nih.gov/clinvar/?term=bmpr2%5Bgene%5D&redir=gene

