## Supplementary figures and images for "Adaptation of ACMG/AMP guidelines for clinical classification of *BMPR2* variants in Pulmonary Arterial Hypertension resolves variants of unclear pathogenicity in ClinVar"

### Supplementary Figure 1

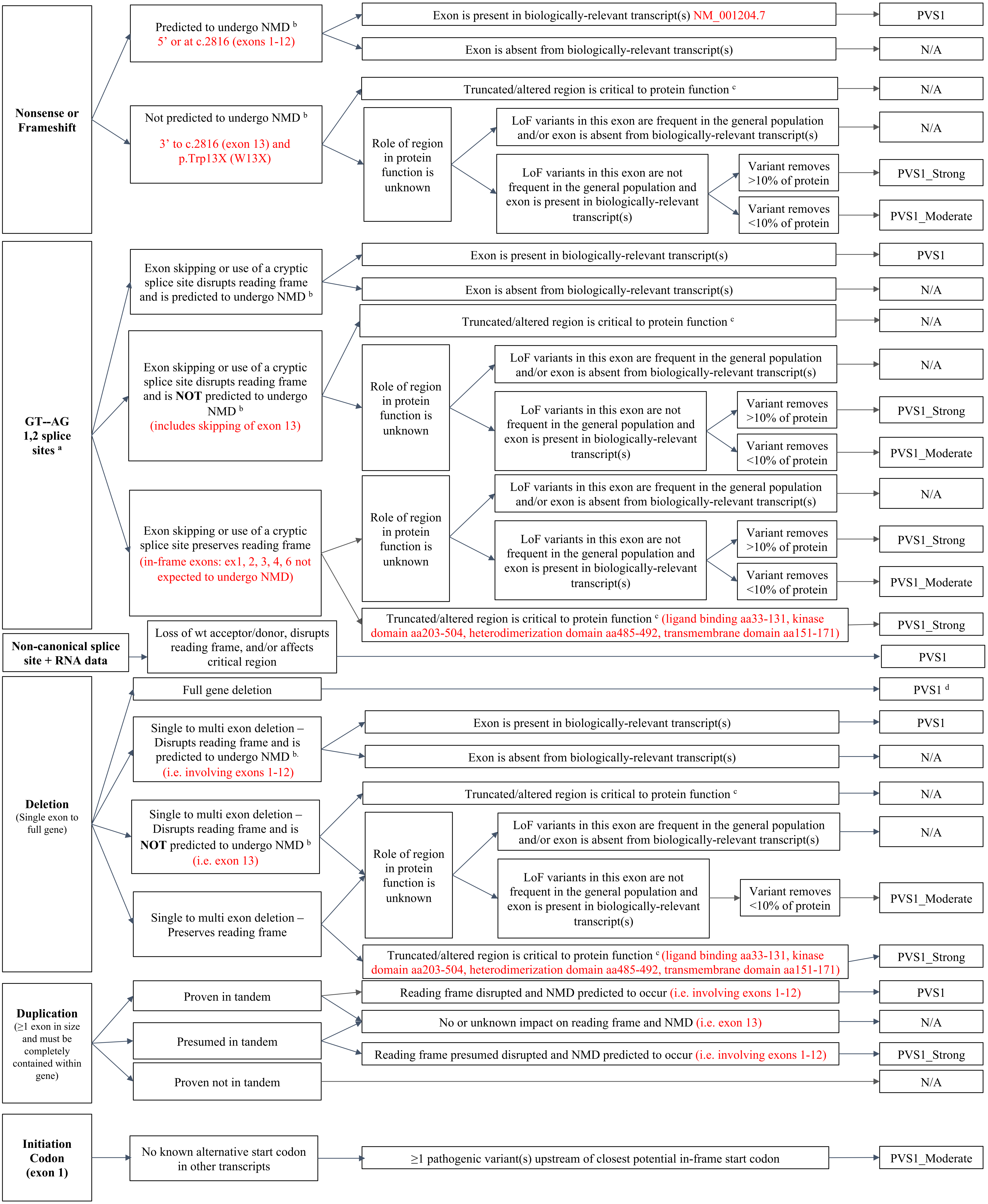
